# The long-term impact and effectiveness of rotavirus vaccination in Malawi: an interrupted time-series and case-control analysis

**DOI:** 10.64898/2026.02.27.26346681

**Authors:** Latif Ndeketa, Virginia E. Pitzer, Khuzwayo C. Jere, Aisleen Bennett, Nigel A. Cunliffe, Peter J. Dodd, Neil French, Daniel Hungerford

**Affiliations:** Institute of Infection, Veterinary and Ecological Sciences, Clinical Infection, Microbiology and Immunology Department, University of Liverpool, Liverpool, UK; Virology Research Group, Malawi-Liverpool-Wellcome Programme, Blantyre, Malawi; School of Life Sciences and Allied Health Professions, Medical Laboratory Sciences Department, Kamuzu University of Health Sciences, Blantyre, Malawi; Department of Epidemiology of Microbial Diseases and Public Health Modeling Unit, Yale School of Public Health, Yale University, USA; National Institute of Health and Care Research Health Protection Research Unit in Gastrointestinal Infections; Sheffield Centre for Health and Related Research, University of Sheffield, Sheffield, UK; Section of Paediatric Infectious Disease, Department of Infectious Disease, Imperial College Healthcare NHS Trust; Imperial College London, London, UK

## Abstract

**Background:** Rotavirus remains a leading cause of childhood diarrhoeal hospitalisation globally. Malawi introduced the monovalent G1P8 rotavirus vaccine (Rotarix®) in October 2012 and in April 2016 switched from trivalent to bivalent oral poliovirus vaccine (tOPV to bOPV). More than a decade after Rotarix® introduction, evidence on sustained vaccine effectiveness and population-level impact in high-transmission, low-income settings remains limited, and it is uncertain whether programme changes such as the OPV formulation switch have influenced Rotarix® performance over time.

**Methods:** We estimated the long-term impact and effectiveness of Rotarix® on diarrhoea hospitalisation in Malawi and explored whether OPV type changes Rotarix® effectiveness. We used interrupted time-series and test-negative case-control analyses to assess the impact and effectiveness of Rotarix®, respectively, over seven years post vaccine introduction using data from diarrhoeal surveillance studies in children under five years hospitalised with acute gastroenteritis at Queen Elizabeth Central Hospital, in Blantyre, Malawi, between 1997 and 2019. Stool samples from these children were collected and tested for rotavirus A antigen using enzyme immunoassay (EIA). To assess the effect of concurrent vaccination with OPV, we used a test-negative case-control study to estimate the interaction between the two vaccines.

**Findings:** The interrupted time-series was based on 7,952 hospitalisations and showed a 23% (95% confidence interval (CI): 10 – 34%) reduction in rotavirus hospitalisations rates among children under five years in the post-vaccine introduction period (January 2013 – December 2019) compared with the pre-vaccine period (July 1997 – December 2012). There was a stronger effect among infants under one year (37%; 95% CI: 25 – 47%). Protection declined with age, with little measurable impact beyond infancy. The test-negative analysis enrolled 1,909 children and Rotarix® effectiveness for two doses was 52% (95% CI: 18 – 71%) overall, 67% (95% CI: 36 – 82%) among infants and 29% (95% CI: −136 – 74%) in those older than one year. Analyses of the tOPV to bOPV switch (n = 1,622) showed no measurable interaction with Rotarix® performance (aOR 1.07; 95% CI: 0.85 – 1.34).

**Interpretation:** Rotarix® provides moderate protection for Malawian infants, and the transition from tOPV to bOPV did not influence vaccine effectiveness. The lower effectiveness of rotavirus vaccination with increasing child age highlights the need to evaluate alternative vaccination strategies alongside strengthened WASH interventions to sustain vaccine impact in LMICs.

**Funding:** MRC Discovery Medicine North (DiMeN) Doctoral Training Partnership (UKRI) and National Institute for Health and Care Research (NIHR)

**Research in context:** *Evidence before this study:* Multiple reviews have evidenced variable vaccine effectiveness by setting and age. A recent global review and meta-regression of efficacy and effectiveness data by the authors (https://doi.org/10.1016/j.eclinm.2025.103122), updated to October 2024, highlighted lower rotavirus vaccine effectiveness and impact in high-burden, low- and middle-income countries (LMICs) compared with high-income settings. Studies in LMIC’s including those in sub-Saharan Africa (SSA) consistently indicate that protection is strongest in infants, with impact and effectiveness declining in older children. Multiple factors have been implicated for this variability in effectiveness, including interference from co-administration of oral polio vaccines. We also conducted a systematic review of post-licensure rotavirus vaccine impact and effectiveness studies from sub-Saharan Africa (CRD42023436851). We also assessed the principal study designs used and the extent to which they adjusted for concurrent public health and social measures (PHSMs). We searched PubMed, EMBASE, MEDLINE, CINAHL, and Google Scholar for studies of vaccine impact or effectiveness in children under five years and screened reference lists of included studies. Across all eligible studies, none measured or adjusted for concurrent public health or social measures. To date, most SSA evaluations have focused on the early post-introduction period, with limited evidence on longer-term vaccine performance and no epidemiological evaluation of the potential effect of co-administration of oral polio vaccines on rotavirus vaccine effectiveness.

*Added value of this study:* We provide a long-term evaluation of the monovalent rotavirus vaccine (Rotarix®) in Malawi using a single hospital-based surveillance platform spanning the pre-vaccine and post-vaccine periods. We combine interrupted time-series analyses of population-level impact with test-negative estimates of individual-level effectiveness using consistent age strata. We also examine whether the national switch from trivalent to bivalent oral poliovirus vaccine modified rotavirus vaccine effectiveness, addressing a programme change that has rarely been assessed in rotavirus vaccine evaluations.

*Implications of all the available evidence:* The available evidence indicates modest rotavirus vaccine benefit in sub-Saharan Africa, with protection concentrated in infancy and little measurable effect in older children. Our findings highlight the need to interpret long-term vaccine impact estimates alongside changes in other PHSMs that influence rotavirus disease burden, including water and sanitation and access to care. The absence of an effect associated with OPV formulation change suggests that modifying OPV valency alone is unlikely to improve rotavirus vaccine performance. Extending protection beyond infancy may require alternative vaccine schedules alongside sustained improvements in broader public health conditions.

## Introduction

Rotavirus infection is the leading cause of severe childhood diarrhoea globally. Prior to the licensure and introduction of rotavirus vaccines, rotavirus was responsible for over 500,000 deaths annually in children under five years of age, with nearly two-thirds of these deaths occurring in low- and middle-income countries (LMICs)[1], [2]. Children in LMICs countries tend to experience rotavirus infection earlier in life when the risk of severe disease is greatest, with peak incidence typically between 9 and 15 months[3]. At the same time these settings face higher case-fatality rates because of limited access to healthcare[4][5].

Four oral live-attenuated licenced rotavirus vaccines are currently recommended by the World Health Organization (WHO). Rotarix® (GlaxoSmithKline Biologicals), RotaTeq® (Merck & Co), Rotavac® (Bharat Biotech) and RotaSiil® (Serum Institute) have been introduced in routine childhood immunisation programmes for over 130 countries [6]. Following introduction, rotavirus vaccines are estimated to have prevented 140,000 deaths from 2006 to 2019[7].

Malawi introduced the Rotarix® vaccine into its Extended Programme for Immunisation (EPI) in October 2012, with a recommended two-dose schedule at 6 weeks and 10 weeks of age. A study conducted two years after vaccine introduction in Malawi estimated a vaccine effectiveness (VE) of 70.6% in children aged <1 year and 31.7% in those aged 1-<2 years[8]. Another study reported 61.89% VE for Rotarix® four years post-introduction in the first year of life[9]. Most recently, Pitzer, *et al* estimated the population-level impact of Rotarix® introduction to be a 36% reduction in the incidence of rotavirus diarrhoea in children <5 years and a 52.5% reduction in children <1 year in Malawi[10].

While these estimates are similar to vaccine efficacy estimated from a phase III randomised clinical trial from Malawi, they remain lower than the VE observed in high income countries HIC[11], [12]. This highlights the need to better understand factors influencing impact and VE in LMICs. One potential factor is the possible interference of oral polio vaccines (OPV) with rotavirus vaccine immunogenicity. Trivalent OPV (tOPV) has been shown to interfere with the immune response to rotavirus vaccines [13]–[15]. This interference is thought to be due to rapid replication of the type 2 poliovirus component of tOPV in the gut at the time of the first tOPV dose, which is administered concurrently with the first Rotarix® dose at 6 weeks of age[16], [17]. Malawi switched from tOPV to a bivalent OPV (bOPV) on 25 April 2016, which only includes type 1 and 3 poliovirus[18]. However, the implications of this switch for rotavirus VE remain unknown.

Our study measures the long-term direct effectiveness (VE) of Rotarix® and impact (overall effectiveness) on rotavirus hospitalisations in children less than 5 years old in Malawi, and the influence of the tOPV to bOPV transition on Rotarix® VE.

## Methods

### Study setting and population

Blantyre is the largest commercial district in the southern region of Malawi. Healthcare is delivered through a network of private and government hospitals, and government health centres. Queen Elizabeth Central Hospital (QECH) is the largest government referral hospital, providing free medical care to the 1.3 million urban, peri-urban and rural residents of Blantyre district.

### Surveillance data

The Malawi Liverpool Wellcome Research Programme (MLW) in partnership with the Ministry of Health established a surveillance programme for children with acute gastroenteritis (AGE) as part of the effectiveness and impact evaluation of Rotarix® following its introduction in the EPI. The surveillance program enrolled children under 5 years old who presented at QECH with diarrhoea lasting less than 14 days and had ≥3 episodes in 24 hours. Stool samples were collected and tested for rotavirus A antigen using enzyme immunoassay (EIA). The surveillance had three periods: July 1997 to June 2007, January 2008 to December 2009, and from January 2012 onwards[19]–[22]. There was a period of no surveillance between December 2009 and November 2011 which was excluded from the analysis. Population denominators were obtained from the National Statistics Office publications of the demographic surveys and their population projections covering the defined period.

### Design

#### Statistical analysis

##### Interrupted time-series to assess long-term vaccine impact

We used an interrupted time-series design to assess the population-level impact of Rotarix® introduction on rotavirus gastroenteritis (RVGE) cases observed from July 1997 to October 2019. We set the change point for the interrupted time-series analysis on 01 January 2013 when Rotarix® vaccine coverage exceeded 80% following the introduction in October 2012. Hospitalisations occurring between October and December 2012 were included in the pre-vaccine period. Monthly aggregates of rotavirus confirmed diarrhoeal admissions were analysed before and after vaccine introduction. To explore changes over time, a fitted time-series model was used to assess changes in both the gradient (slope) and level (intercept) of diarrhoeal cases post-vaccine introduction. We used generalized linear regression models with negative binomial distributions to account for the overdispersion in the data. The models included terms for seasonality (seasonal harmonics) and secular trends (linear year term).

To adjust for changes in healthcare seeking and surveillance intensity over time, we included test-negative diarrhoeal cases as a proxy for background admission trends independent of vaccination. Yearly census data for all the age categories were obtained from the National Statistics Office of Malawi and provided a robust population offset for the analysis. The vaccine impact on the hospitalisation rate was calculated as *100 X (1 - Incidience Rate Ratio*) with associated 95% confidence intervals (95% CIs)

For the counterfactual analysis, we used our primary analytic model to predict hospitalisation trends in the absence of vaccination. The factual series reflected observed vaccination effects while the counterfactual series was generated by setting the vaccine indicator variable to zero and keeping all other covariates unchanged. Monthly observed case counts were visualised alongside the factual and counterfactual predictions with solid and dashed lines used to distinguish the two. We performed all analyses separately for children aged <1 year, ≥1 to <5 years, and <5 years.

##### Test-negative case-control study for vaccine effectiveness

We conducted a test-negative case-control study to evaluate Rotarix® effectiveness, comparing the odds of vaccination among cases and controls from October 2012 to December 2019. We included children born on or after 17 September 2012 as they would have been 6 weeks old when the vaccine was introduced and eligible for vaccination for the first Rotarix® dose. Children were classified as vaccinated only if they had completed the full two-dose schedule. For consistency with the interrupted time-series analysis, we estimated vaccine impact separately for children aged <1 year, ≥1 to <5 years, and <5 years.

Vaccine status was obtained through hand-held patient medical records (health passports). Children who were verbally reported to be vaccinated but had no record in the health passport were excluded from the analysis. Cases were defined as children who presented with AGE and tested rotavirus EIA positive (RVGE), while controls were children who presented with AGE and tested rotavirus EIA negative. We measured the odds of vaccination between cases and controls adjusting for sex, age at admission, and month and year of birth using logistic regression. Vaccine effectiveness was calculated as (1 *- Odds Ratio) X* 100% and presented with 95% CIs.

##### Relative effectiveness of the switch from tOPV to bOPV on Rotarix®

A test-negative case-control design was used to assess the relative VE of Rotarix® following the switch from tOPV to bOPV. Specifically, we analysed if children vaccinated with dose one of bOPV (administered at 6 weeks concurrently with Rotarix®) were at a lower risk of laboratory-confirmed rotavirus hospitalisation than children vaccinated with tOPV and Rotarix®. The analysis utilised a subset of the same surveillance data previously described and covered the post-Rotarix® introduction period between January 2013 to December 2019. Only children who received both Rotarix® and OPV were included in the analysis.

The exposure variable was vaccination with bOPV and included all infants who were vaccinated with oral polio vaccine after 31^st^ May 2016. The unexposed group represented infants who received their oral polio vaccine before 1^st^ of April 2016 (tOPV vaccinated). The months of April and May 2016 were excluded from the analyses to reduce the risk of cross-contamination in the exposure groups related to lags in vaccine switch. The adjusted odds ratio (aOR) was calculated using logistic regression with adjustments for age at admission, month of birth, sex and nutritional status measured by mid upper arm circumference (MUAC). Vaccine effectiveness was calculated as (1 *- aOR) X* 100% *and pres*ented with 95% CIs. All statistical analyses were performed in R Version 4.2.3.

## Results

The interrupted time-series analysis examined 7,952 AGE cases across the periods before and after Rotarix® introduction. The test-negative case-control study included 1,909 children, from which a subset of 1,622 was used for the OPV-Rotarix® interaction analysis. We observed a shift in the age distribution of rotavirus-confirmed hospitalisations towards older children following Rotarix® introduction. In children aged ≥12 months, the mean annual number of RVGE hospitalisations increased from 22.7 cases per year in the pre-vaccine period to 51.3 cases per year after January 2013. Over the same period, mean annual RVGE hospitalisations in infants aged <12 months declined modestly, from 86.8 to 76.6 cases per year. These patterns indicate that, while RVGE burden in infancy decreased following Rotarix® introduction, a greater share of hospitalised disease occurred among older children.

Figure 1 illustrates the modelled trend of rotavirus-positive cases before vaccine introduction extended to show the expected pattern had Rotarix® not been introduced (counterfactual). The monthly trend of rotavirus-positive and negative AGE cases before and after Rotarix® introduction among children <5 years is shown in Supplementary Figure S1.

**Figure 1.**
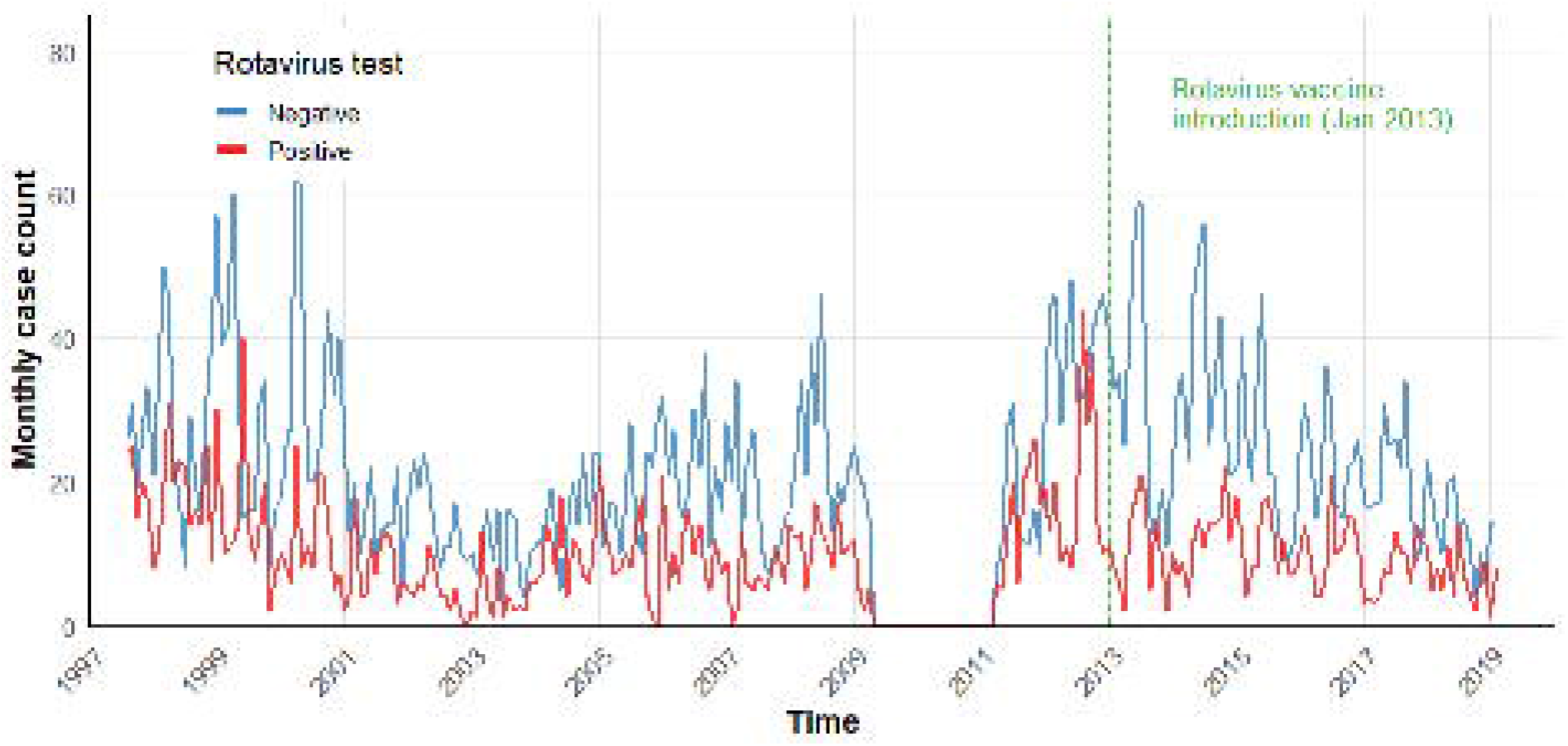
Observed versus counterfactual RVGE cases before and after vaccine introduction. Change point was set on January 2013 but the vaccine was introduced in October 2012

Our analysis revealed age-specific patterns in Rotarix® impact on hospitalisations. There was an absolute 3.5% reduction in RVGE prevalence among children aged <5 years (from 33.2% to 29.7%) and an overall vaccine impact of 23% (95% CI: 10 – 34%, p = 0.001) (Table 1). Among infants <1-year, post-vaccine RVGE prevalence was reduced by 9.7% (from 39.3% to 30.4%), corresponding to a 37% vaccine impact (95% CI: 25 – 47%, p<0.001). In contrast, we observed an increased burden of RVGE to older age groups; RVGE prevalence increased by 7.5% in children ≥1 to 5 years old (from 23.9% to 31.4%) with a non-significant effect estimate of -39% (95% CI: -70 – 24%, p=0.52) (Table 1).

**Table 1.** Negative binomial regression results showing the impact of Rotarix® introduction on rotavirus-positive hospitalisations by age group, adjusted for seasonality, secular trends, and test-negative controls.

| Age group | Pre-vaccine RVGE prevalence | Post-vaccine RVGE prevalence | Adjusted IRR (95% CI) | Vaccine Impact % (95% CI) | P-value |
| --- | --- | --- | --- | --- | --- |
| <5 years | 33.2% (1,643/4,944) | 29.7% (895/3,008) | 0.77 (0.66 – 0.90) | 23 (10 – 34) | 0.001 |
| <1 year | 39.3% (1,302/3,310) | 30.4% (536/1,764) | 0.63 (0.53 – 0.75) | 37 (25 – 47) | <0.001 |
| $\geq 1$ year | 23.9% (304/1273) | 31.4% (310/987) | 1.39 (1.13 – 1.70) | -39 (-70 – -13) | 0.002 |

### Variation in Rotarix® impact over time

Yearly estimates across seven years after Rotarix® introduction also showed age□dependent variation in impact, but vaccine impact remained fairly consistent over time (Figure 2). Among children <5 years, the overall vaccine impact was modest, with only the second year after introduction showing a significant reduction in RVGE hospitalisations . The IRR in children aged <1 year varied between 0.56 and 0.74 and was highest in 2016. The IRR >1 among children aged ≥1 to <5 years, indicating that an increased incidence in older children mostly occurred right after vaccine introduction. More detailed results are presented in the supplementary table S1.

**Figure 2.**
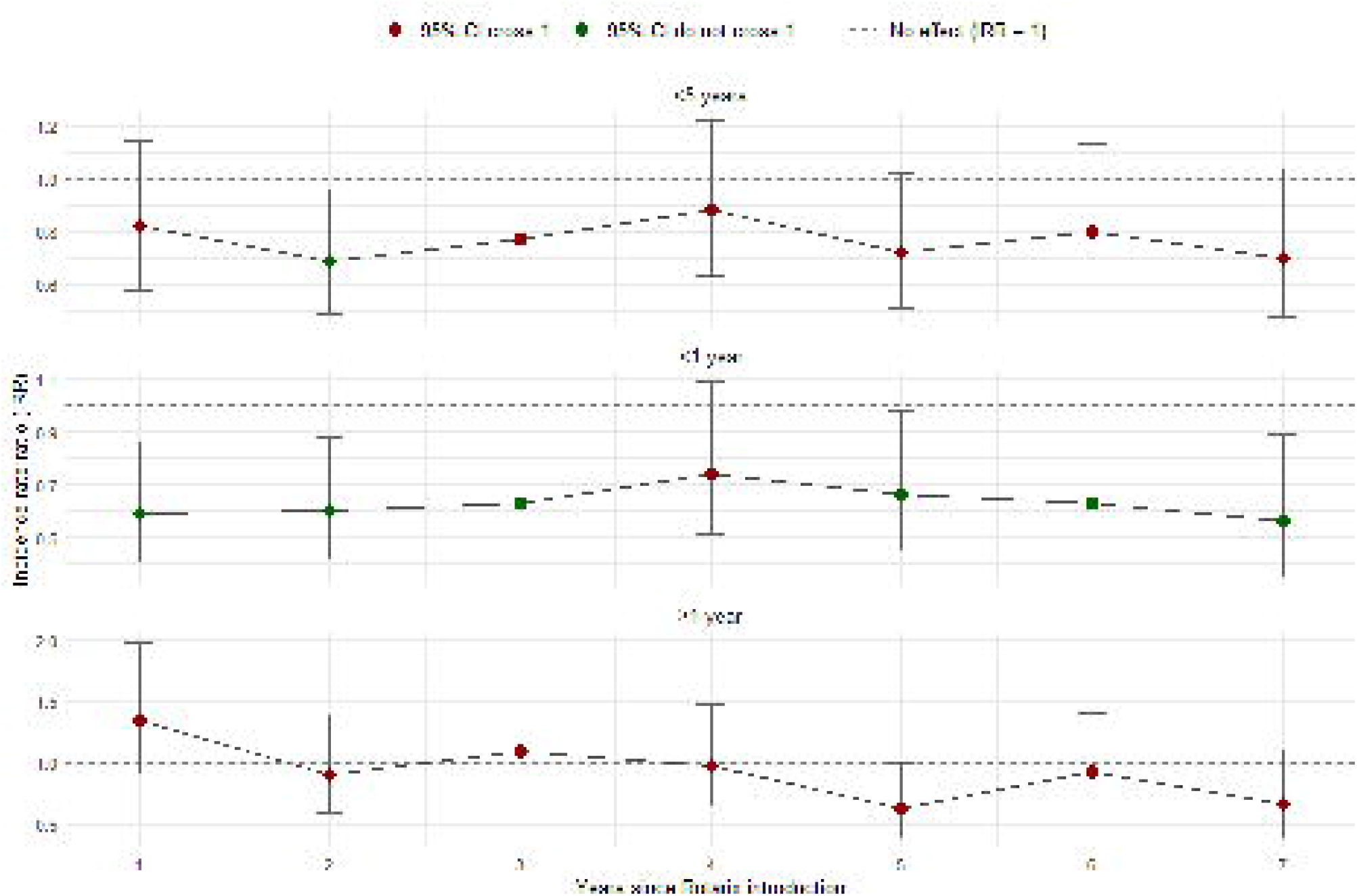
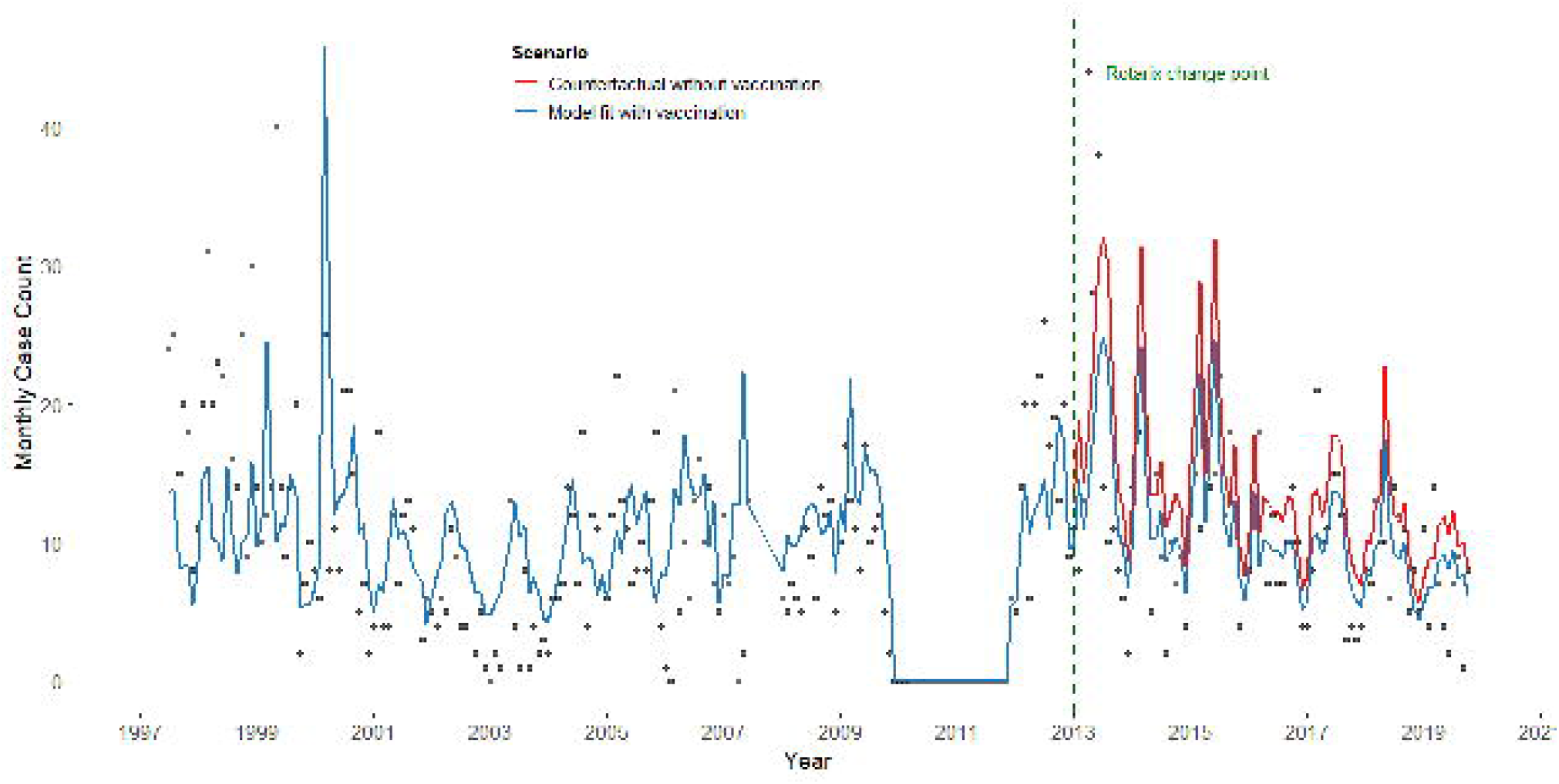
Annual estimates of rotavirus vaccine impact on RVGE hospitalisations following Rotarix® introduction by age group. Points show adjusted incidence rate ratios (IRRs). Error bars represent 95% confidence intervals

### Vaccine effectiveness

The test-negative case-control study included 1,909 children hospitalized with AGE between January 2013 and December 2019, comprising 551 rotavirus-positive cases (28.9%) and 1,358 test-negative controls (71.1%). The median age was 10.6 months (interquartile range: 8 - 14.8 months), with 64.4% of cases occurring in infants <1 year. More details on the baseline characteristics are presented in Supplementary Table S3. Complete two-dose Rotarix® vaccination demonstrated 52% protection against rotavirus hospitalization in children <5 years (aOR 0.45, 95% CI 0.29-0.82; p = 0.006).

Vaccine effectiveness differed across age groups as shown in Table 2. Among children <1 year, protection reached 67% (aOR 0.33; 95% CI 0.18–0.64; p = 0.001). Vaccine effectiveness in children older than 1 year was lower at 29% (aOR 0.78; 95% CI 0.26–2.32; p = 0.50).

**Table 2.** Regression estimates and 95% confidence intervals for models used to estimate Rotarix® vaccine impact and effectiveness across age strata.

| Age Group | Cases/Controls | Unadjusted OR (95% CI) | Adjusted OR† (95% CI) | Vaccine Effectiveness‡ (95% CI) | P-value |
| --- | --- | --- | --- | --- | --- |
| <5 Years | 551/1,358 | 0.49 (0.30–0.81) | 0.48 (0.29–0.82) | 52% (18–71) | 0.006 |
| <1 year | 355/822 | 0.40 (0.22–0.73) | 0.33 (0.18–0.64) | 67% (36–82) | 0.001 |
| >1–<5 years | 196/536 | 0.83 (0.33–2.43) | 0.78 (0.26–2.32) | 29% (–136 to 74) | 0.500 |

†Adjusted for age (months), birth month, birth year
‡Calculated as $(1 - \text{aOR}) \times 100\%$

### Interaction of OPV and Rotarix® vaccination

The analysis included 1,622 children who received both OPV and Rotarix®, with 504 rotavirus cases and 1,118 test-negative controls. The proportion receiving bOPV was similar between cases (32.3%) and controls (30.3%) (Table 1). Age, sex and nutritional status were evenly distributed across the two groups, and most participants were <1 year of age (64.7% of cases and 65.0% of controls). More details of the descriptive statistics are presented in Supplementary Table S2.

In our crude analyses, the odds of laboratory-confirmed rotavirus hospitalisation were similar among children who received bOPV alongside Rotarix® and those who received tOPV alongside Rotarix® (OR 0.87; 95% CI 0.70–1.08; p = 0.21). We found no evidence of an association between OPV formulation and Rotarix® vaccine effectiveness (Table 3). After adjusting for age, sex and nutritional status, the association remained non-significant (aOR 1.07; 95% CI 0.85– 1.34; p = 0.58). Figure 6 illustrates this lack of measurable difference in Rotarix® effectiveness between children vaccinated with bOPV versus tOPV. Children with severe acute malnutrition had significantly lower odds of RVGE compared with well-nourished peers (aOR 0.60, 95% CI 0.42–0.85; p=0.005), whereas no association was observed among those with moderate malnutrition.

**Table 3.**
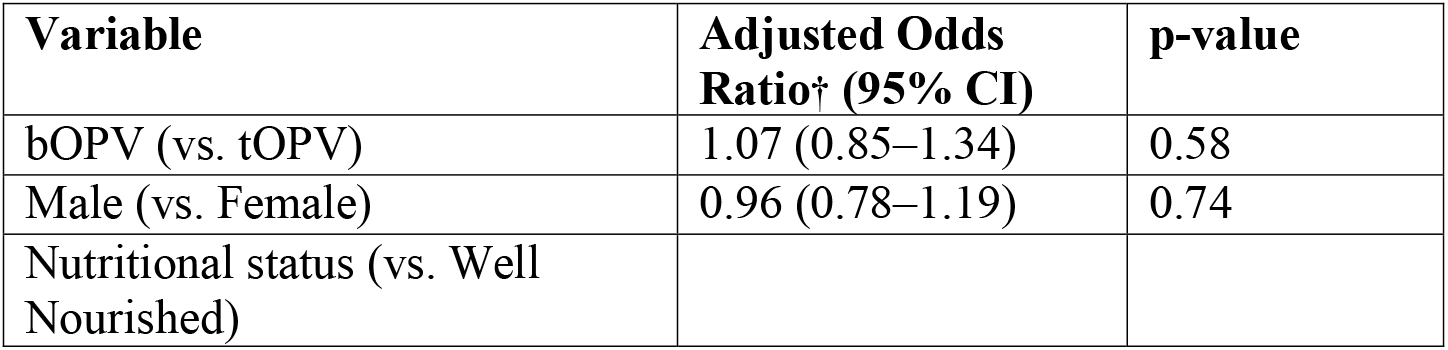
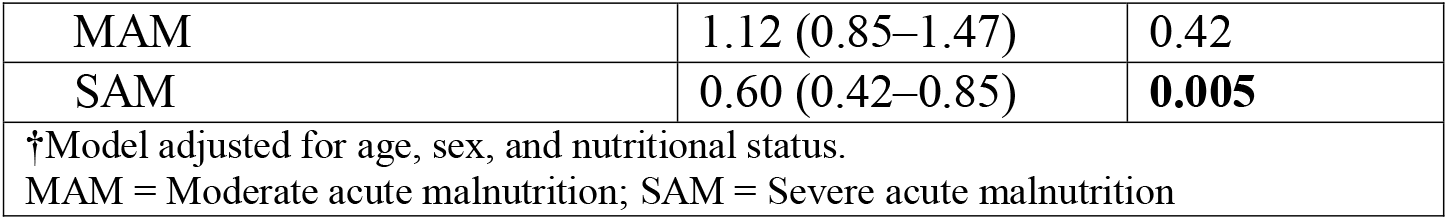
Results of the interaction analysis between oral poliovirus vaccine (OPV) and rotavirus vaccine (Rotarix®) showing adjusted estimates of Rotarix® effectiveness among children eligible for both vaccines.

## Discussion

Rotarix® continues to provide modest protection against RVGE hospitalisation in Malawi more than seven years after its introduction. Vaccine effectiveness and impact analyses showed sustained benefit in infancy, with protection highest in the first year of life. Protection declined with age, and among children >1 year neither analysis demonstrated clear benefit, with effectiveness estimates overlapping the null and impact estimates indicating increased admissions.

We observed a shift in the age distribution of rotavirus-confirmed hospitalisations towards older children following Rotarix® introduction. Mean annual RVGE hospitalisations among children aged ≥12 months increased in the post-vaccine period, while admissions in infants declined modestly, indicating that protection was concentrated in the first year of life. This pattern likely reflects delayed primary infection alongside waning vaccine-derived protection, with child age acting as a proxy for time since vaccination. The year 1 estimate in children aged ≥1 year is difficult to attribute to vaccination, since most children in this stratum would not have been vaccine eligible during the initial rollout period. Consistent with this interpretation, we observed no evidence that the switch from tOPV to bOPV influenced Rotarix® effectiveness in this population.

Our findings align with those of Pitzer *et al*., who estimated the impact of Rotarix® in Malawian children to be 36% (95 % prediction interval 33.6–39.9) in those <5 years and 52.5% (95% prediction interval 50.1–54.9) in infants <1 year[10]. The authors used validated transmission models to predict the incidence of RVGE hospitalisation over the 10-year period following vaccine introduction. Here, we took a more traditional interrupted time-series-based approach to predict the counterfactual incidence of RVGE in the absence of vaccination. Our interrupted time-series analysis partially controlled for the effect of long-term underlying trends which could potentially explain the decrease in rotavirus hospitalisations due to improved access to water, sanitation and hygiene (WASH) over the analysis time period if it coincided with the introduction of Rotarix® in Malawi[23]. Our model also adjusted for time-varying confounders and year-to-year seasonal variation of the RVGE cases over time. We included rotavirus test-negative cases as a covariate in our analysis to adjust for any effect on the vaccine impact that could be introduced by the changes in surveillance methods over time, differences in healthcare seeking behaviour among children and any improvements in WASH interventions over time.

Both studies indicated reduced or negative impact estimates in children older than 1 year. The observed upward age shift in RVGE epidemiology is commonly observed in vaccination programmes and is largely attributed to waning of vaccine-induced immunity and delayed exposure to primary infection following high vaccination coverage [24]. Our findings indicate that protection declines beyond infancy suggesting the need for a booster dose of Rotarix® to sustain protection in older children, which aligns with other studies on Rotarix® performance in Malawi [25], [26]. Mandolo *et al*. recommend 6 to 8 months as the optimal window for administering a booster in an LMIC, high-disease-burden setting[27]. Whilst our results support the provision of a booster in children >1 years, policy implementers must consider the programmatic feasibility and cost-effectiveness of a booster dose. Previous analyses suggest the provision of an additional dose at 14 weeks of age may be cost-effective in Malawi and could be integrated within the EPI schedule[28]. A further dose administered at 14 weeks of age or nine months concurrently with the measles vaccine could be considered to strengthen rotavirus protection without requiring an extra visit on the immunisation schedule [29], [30].

We found no evidence that the switch from tOPV to bOPV altered the effectiveness of Rotarix®. This aligns with immunogenicity data from Bangladesh showing reduced Rotarix® seroconversion when Rotarix® and OPV are administered on the same day, with similar interference observed across monovalent, bivalent, and trivalent OPV formulations[15]. These findings suggest that changes in OPV formulation alone may not remove oral vaccine interference. Other mechanisms may contribute to reduced oral vaccine performance in low-income settings. For example, higher gut microbiota diversity in early infancy has been associated with lower Rotarix® seroconversion in several LMIC studies [31] suggesting that biological and environmental factors beyond OPV formulation remain important. The association we observed between severe malnutrition and RVGE hospitalisation should also be interpreted cautiously, as similar studies in India, Nigeria, and Pakistan have not reported consistent evidence of this relationship[32]–[34].

A neonatal schedule could also be an alternative to a booster dose to optimise protection in the face of interference from other oral vaccines. RV3-BB, a neonatal strain rotavirus vaccine (G3P[6]) currently in Phase III trials, has shown improved replication in the newborn gut and may be less affected by maternal antibodies, potentially offering an effective option for more durable protection [35], [36]. Use of neonatal vaccine has been predicted to be the most cost-effective option for Malawi in a recent modelling study [28]. This is supported by clinical data showing that the neonatal rotavirus vaccine RV3-BB is safe and immunogenic when administered at birth or in early infancy in Malawi, with serological responses comparable to those achieved with current schedules[37].

There were several limitations to our data and analysis. Although our surveillance data was robust, vaccine coverage data for Blantyre were unavailable to include in our negative binomial model for vaccine impact. Rotarix® coverage has exceeded 80% since January 2013, so calendar time was used as a proxy for vaccine exposure, which is likely to be a reasonable approximation in this setting[38]. Finally, QECH caters to the Blantyre urban catchment area, where improved WASH may lead to a higher estimated vaccine impact, resembling what is observed in high-income countries with strong WASH infrastructure. As a result, impact estimates from this analysis may be higher than those that would be observed in more rural parts of Blantyre, where poorer baseline WASH conditions may sustain a higher underlying burden of disease. In addition, confidence intervals around vaccine effectiveness estimates in children aged >1 to <5 years were wide, reflecting limited precision as RVGE admissions declined in older age groups.

## Conclusion

Our study provides a comprehensive evaluation of Rotarix®’s long-term effectiveness and impact in Blantyre, Malawi. This analysis demonstrated strong protection among infants. Vaccine protection declined with age and led to a gradual shift in disease burden towards older children. Despite these shifts, Rotarix® maintained a significant overall impact reducing RVGE incidence in the vaccine-eligible population. The transition from tOPV to bOPV did not appear to influence vaccine effectiveness, suggesting minimal interference with the immune response. Evaluation of neonatal vaccination or booster dosing alongside strengthened WASH interventions should be prioritised to sustain vaccine impact in LMICs.

## Data Availability

All data produced in the present study are available upon reasonable request to the authors
All data produced in the present work are contained in the manuscript

## Contributions

L.N., D.H., N.F, N.A.C, and K,C.J conceived of the study and secured the funding. L.N. performed the analysis, wrote the manuscript. N.F., V.E.P, D.H., P.D assisted with the analysis and interpretation of results and edited the manuscript. K.C.J. A.B., and N.A.C. oversaw the data collection. All authors assisted with the interpretation of results and edited the manuscript.

