## Supplementaty tables and figure for "The long-term impact and effectiveness of rotavirus vaccination in Malawi: an interrupted time-series and case-control analysis"

**Supplementary**

**Supplementary Table S1.** Annual variation in rotavirus vaccine effectiveness (Rotarix®) by age group since introduction showing trends in protection among children under five years of age

| **Years post vaccine introduction** | **<5 IRR (95% CI)** | **<1 IRR (95% CI)** | ≥**1 IRR (95% CI)** |
| --- | --- | --- | --- |
| 1 | 0.82 (0.58–1.14) | 0.59 (0.41 – 0.86)****** | 1.35 (0.92–1.98) |
| 2 | 0.69 (0.49–0.96) * | 0.60 (0.42 – 0.88)****** | 0.91 (0.59–1.39) |
| 3 | 0.77 (0.55–1.08) | 0.63 (0.44 – 0.92)***** | 1.10 (0.73–1.63) |
| 4 | 0.88 (0.63–1.22) | 0.74 (0.51 – 1.09) | 0.98 (0.65–1.48) |
| 5 | 0.72 (0.51–1.02) | 0.66 (0.45 – 0.98)***** | 0.63 (0.39–1.00) |
| 6 | 0.80 (0.57–1.13) | 0.63 (0.42 – 0.93)****** | 0.93 (0.61–1.40) |
| 7 | 0.70 (0.48–1.04) | 0.56 (0.35 – 0.89)***** | 0.67 (0.39–1.11) |
| ***<0.05, **<0.01, ***<0.001** | | | |

**Supplementary Table S2**: Characteristics of rotavirus-positive cases and test-negative controls included in the OPV–Rotarix® interaction analysis (children who received both OPV and Rotarix®)

| **Characteristic** | **Controls (N=1,118)** | **Cases (N=504)** |
| --- | --- | --- |
| Received bOPV, n (%) | 339 (30.3%) | 163 (32.3%) |
| Age category, n (%) |  |  |
| <1 year | 727 (65.0%) | 322 (63.9%) |
| 1–<2 years | 332 (29.7%) | 163 (32.3%) |
| ≥2 years | 59 (5.3%) | 19 (3.8%) |
| Male sex, n (%) | 693 (62.0%) | 310 (61.5%) |
| Nutritional status, n (%) |  |  |
| MAM | 200 (17.9%) | 105 (20.8%) |
| SAM | 151 (13.5%) | 43 (8.5%) |
| Well-nourished | 766 (68.6%) | 356 (70.6%) |
| MAM = Moderate acute malnutrition; SAM = Severe acute malnutrition | |  |

**Supplementary Table S3:** Characteristics of rotavirus-positive cases and rotavirus-negative controls

| **Characteristic** | **Controls (N=1,118)** | **Cases (N=504)** |
| --- | --- | --- |
| **Received bOPV, n (%)** | **339 (30.3%)** | **163 (32.3%)** |
| **Age category, n (%)** |  |  |
| **<1 year** | **727 (65.0%)** | **322 (63.9%)** |
| **1–<2 years** | **332 (29.7%)** | **163 (32.3%)** |
| **≥2 years** | **59 (5.3%)** | **19 (3.8%)** |
| **Male sex, n (%)** | **693 (62.0%)** | **310 (61.5%)** |
| **Nutritional status, n (%)** |  |  |
| **MAM** | **200 (17.9%)** | **105 (20.8%)** |
| **SAM** | **151 (13.5%)** | **43 (8.5%)** |
| **Well-nourished** | **766 (68.6%)** | **356 (70.6%)** |
| **MAM = Moderate acute malnutrition; SAM = Severe acute malnutrition** | |  |

**Supplementary Figure 1.** Monthly trend of rotavirus positive and negative acute gastroenteritis (AGE) cases before and after Rotarix® introduction among children under five years

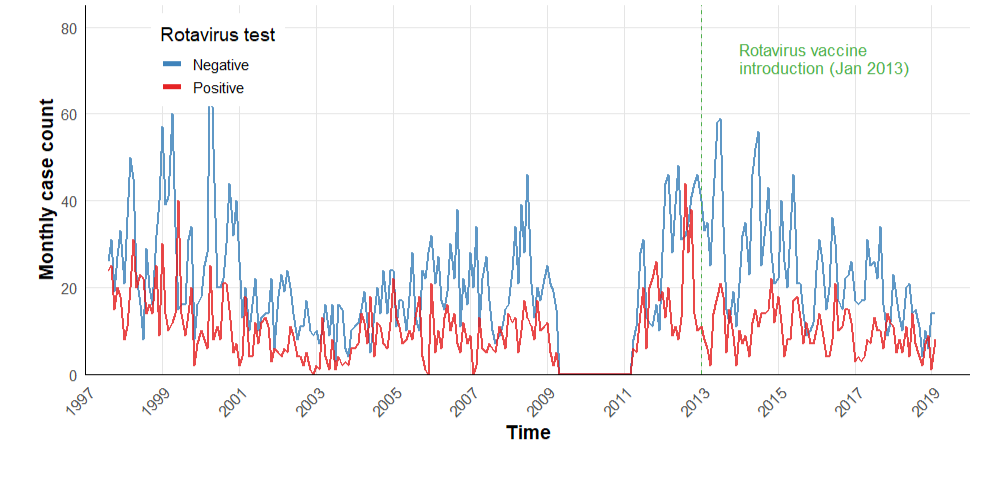
